# Hypothesis-driven patterns of intracranial exploration in temporal lobe epilepsies

**DOI:** 10.1101/2024.07.21.24310240

**Authors:** Arka N. Mallela, Jasmine Hect, Eliza Reedy, Naoki Ikegaya, Hussam Abou-Al-Shaar, Theodora Constantine MPAS, PA-C, Arthur Angonese, Thandar Aung, Luke C. Henry, Danielle R. Carns, Jorge A. González-Martínez

## Abstract

**Background:** Surgical treatment is a highly effective option for drug-resistant temporal lobe epilepsy (TLE). When non-invasive exploration is insufficient to localize the epileptogenic zone, anatomo- electroclinical (AEC) hypotheses can be interrogated by stereoelectroencephalography (SEEG) methodology. To facilitate more effective exploration and intervention, we developed a classification system of AEC hypotheses for temporal lobe SEEG explorations: Mesial lateral temporal (MLT), temporal basal occipital (TBO), anterior perisylvian (AP), and perisylvian (P).

**Methods:** In a cohort of 60 patients with drug-resistant TLE who underwent SEEG, we collected demographic data, clinical/epilepsy history, neuroimaging, seizure semiology, and the analysis of the multidisciplinary epilepsy patient management conference. We identify and describe the 4 patterns of hypotheses, analyze the semiological and structural features, and validate the proposed classification system using a random forest classifier machine learning algorithm.

**Findings:** Using this approach, we identify the features most predictive of each classification, and demonstrate an overall 90% classification accuracy using semiological features and 97% accuracy using electrode location. Age, sex, and the presence of an MRI abnormality did not vary by classification. We qualitatively explain the features of each classification with case examples. Finally, we specifically provide indirect targeting coordinates for each electrode to facilitate reproducible SEEG explorations. Following SEEG exploration, 94% patients underwent surgical intervention (82% selective resection, 12% neuromodulation). In resection patients, one-year seizure freedom rate was 73% and did not vary significantly by classification (MLT: 86%, TBO: 71%, AP: 75%, P: 68%; p=0.9) with overall 26% verbal memory decline.

**Interpretation:** The patterns of explorations, grounded in semiology and cortical cytoarchitectonic and functional features, guided volumetrically-restricted resections that resulted in a high rate of seizure freedom. This approach guides both a theoretical approach to TLE and a practical realization that can be tailored to the individual patient.

**Research in context:** *Evidence before this study:* We searched the MEDLINE database from inception to June 1, 2024 using the terms (“temporal lobe epilepsy”, “mesial temporal lobe epilepsy”, “anatomo-electroclinical hypothesis”, “stereo EEG”, “neuropsychological outcome”, “surgical strategy”, “intracranial exploration”, and/or “seizure freedom”. We supplemented this with search of Google Scholar and reference list. After review, we selected 1 randomized controlled trial (RCT), 17 cohort studies, 9 technical reports, 1 epidemiological report, and 2 systematic reviews. The RCT compared surgical treatment of temporal lobe epilepsy vs. medical management using a standardized temporal lobectomy approach in all patients and demonstrated significant benefit for surgical treatment. However, it did not address the process of anatomo-electroclinical hypotheses to guide intracranial exploration and selective temporal resections nor did it address neuropsychological outcomes. The cohort studies varied in focus, but described various semiological features, surgical approaches, and neuropsychological outcomes. Despite these reports, a systematic approach to intracranial exploration using SEEG and detailed analysis of seizure freedom and neuropsychological outcomes is lacking.

**Added value of this study:** Our study addresses this limitation in one of the largest cohorts of TLE patients with SEEG explorations to date. Using semiological observations and a detailed cytoarchitectonic analysis, we propose a novel hypothesis classification system of four hypotheses in TLE and describe this in detail. We quantitatively validate this approach using a machine learning based approach and provide details to facilitate a practical implementation. Finally, we demonstrate a high rate of seizure freedom with excellent neuropsychological outcomes using this approach.

**Implications of all of the available evidence:** In this study, we demonstrate that that an appropriately designed intracranial exploration (based on the proposed hypothesis classification) can provide excellent results without necessitating a standard temporal lobectomy. This challenges the prior selective vs. standard temporal dichotomy in the literature but builds on several themes previously reported. This precision and patient-centered approach integrates multimodal information and multidisciplinary discussion resulting in more selective surgical interventions that maximize seizure freedom while minimizing neuropsychological morbidity.

## Introduction

Drug-resistant temporal lobe epilepsy (TLE) is the most prevalent form of refractory epilepsy, imposing substantial neurological, psychological, and financial burdens, and frequently necessitating surgical evaluation^1^. Epilepsy surgery aims to stop seizures by treating the cortical regions responsible for the primary organization of the epileptogenic activity, known as the epileptogenic zone (EZ)^2–4^. The EZ can overlap with anatomical regions associated with brain function. Thus, preservation of these regions is another goal of surgical resection. The assessment of the EZ depends on the formulation of specific anatomo-electroclinical (AEC) hypotheses that consider the temporal-spatial patterns of ictal events, using seizure semiology, patient anatomy, and electrophysiological data^5^.

In stereoelectroencephalography (SEEG), precise electrode placement guided by these hypotheses allows electrophysiological exploration of cortical and subcortical structures. To accurately interrogate the AEC, *in situ* recording of ictal and interictal activity is essential. Thus, appropriate placement of electrodes is fundamental. To achieve reproducibility and precision, stereotaxic coordinate systems based on fixed cerebral landmarks such as the anterior and posterior commissures have been described and validated. These systems facilitate precise indirect targeting and enhance the consistency of SEEG implantations to explore specific hypotheses of localization^6–10^.

Hypotheses in TLE regarding localization have been stratified according to diverse clinical and electrophysiological criteria. Previous classification frameworks predominantly segregated TLE into two main categories: mesial TLE, primarily encompassing the hippocampus, entorhinal cortex, and amygdala, versus lateral or neocortical TLE^9,11–13^. Furthermore, surgical interventions for TLE have traditionally been categorized into selective approaches, predominantly targeting mesial structures^14,15^, including laser interstitial thermal therapy^16,17^, and standard temporal lobectomies involving resection of all lateral temporal structures to approximately 4-4·5cm from the temporal pole on the dominant side and 5-6cm on the non-dominant side, coupled with mesial structures^18^.

However, the dichotomy of mesial/lateral classification often amalgamates cortical regions with markedly distinct properties and separates regions that are intricately interconnected^19^, thereby necessitating a classification schema that duly acknowledges the diversity of regions within and interconnected to the temporal lobe. This classification system aims to optimize intracranial implantation strategy by bridging commonly observed patterns of seizure dynamics to individual semiological and anatomic nuances to enhance the precision and effectiveness of epilepsy surgical treatment.

To elucidate and provide empirical grounding for this approach to SEEG exploration, we analyzed AEC hypotheses in a cohort of 60 drug-resistant TLE patients who underwent SEEG. Drawing from insights gleaned from this cohort and our accumulated expertise from over 850 previous SEEG patients, we formulate a classification framework comprising four distinct subtypes for TLE hypotheses and specify SEEG implantation strategies to explore each hypothesis, including case illustrations. We rigorously validate this taxonomy using a machine learning methodology to compare the classification to seizure semiology and cytoarchitectonic regions. Finally, utilizing this approach, we demonstrate a high rate of seizure freedom and neuropsychological preservation with appropriately tailored selective resections.

## Methods

### Patient selection and data collection

This study was approved by our institution’s IRB (STUDY #21020058). The STROBE guidelines were followed in the preparation of this manuscript. For this report, we only considered the orthogonally oriented trajectories due to the consistency of implantation and the simultaneous exploration of the mesial and lateral compartments in the temporal lobe. We prospectively screened 77 patients who underwent SEEG for drug-resistant epilepsy between 2019-2022 at our institution. Inclusion criteria were adult patients (≥18 years) and hypotheses related to the temporal lobe and adjacent cortical regions. Exclusion criteria were insufficient (<1 year) follow-up period after SEEG-guided resection. Sixty patients were selected.

Demographics, clinical/epilepsy history, and neuroimaging were collected from the electronic medical records. Seizure semiology was collected from the history of the treating epileptologist, interpretation of the epilepsy monitoring unit epileptologist during the patient’s video EEG monitoring, interpretation of the multidisciplinary epilepsy conference, and review of video EEG by the authors. Seizure outcomes was determined from the electronic medical records and confirmed with in person clinical assessment with seizure freedom defined as Engel I. A clinically significant surgical complication was defined as any unexpected event that occurred during SEEG placement, extraoperative monitoring, or removal that resulted in a change in clinical care or neurological status. The details of phase I evaluation, neuropsychological testing, preoperative planning and surgical technique, electrode trajectories (**Supplementary** Figure 1-3), electrode reconstruction, and resection segmentation are described in the Supplementary Methods.

### Hypothesis classification and case examples

The authors defined four hypothesis classifications *a priori* for didactic clarity and to provide a schema for implantations. For each patient, the authors retrospectively reviewed the patient’s multidisciplinary epilepsy patient management conference notes, SEEG implantation scheme, and medical records with a detailed reviewed of ictal semiology captured by video-EEG monitoring sessions. We focused on the main ictal semiological and structural features from the studied cohort. Other factors including structural and functional imaging and Phase I electrophysiology were also taken into consideration for planning the implantation of electrodes. Classification was confirmed by consensus among the authors. To optimally illustrate the classification subtypes, case examples were selected and discussed in detail.

### Feature selection and machine learning classification

To quantitively validate our proposed classification over electrode location, semiology, and resection regions, we utilized random forest (Gini criterion, 100 trees, 1 sample per leaf, 2 samples per split)-based recursive feature elimination (RFE) to identify the 10 features with the greatest impact on classification prediction accuracy. Briefly, this approach begins with entire set of features and eliminates those with the least predictive accuracy until the desired number of features is obtained.

To then validate the combination of these 10 features we used a second random forests classifier (Gini criterion, 500 trees, 1 sample per leaf, 2 samples per split) to determine overall classifier accuracy. Further details of machine learning and statistical analysis are described in the Supplementary Methods.

## Results

### Cohort characteristics

The study cohort consisted of 60 patients with drug-resistant TLE who were referred for invasive Phase II monitoring. Demographic and surgical details are reported in **Table 1**. The cohort had a median age of 34 [28, 44] years at the time of SEEG and were 55% male. Age and sex did not vary significantly by hypothesis (p=0.3 for both). The median number of SEEG electrodes was 14 [13, 15] and was significantly different (p=0·044) between classifications. Only 1 patient of 60 (1·7%) had additional electrodes implanted due to insufficient coverage of the EZ. The overall clinically significant complication rate from SEEG was 3·3% (2/60) and did not vary by hypothesis classification (p=0·8).

**Table 1:** Cohort characteristics by hypothesis

| Characteristic | Overall,<br>N = 60<sup>1</sup> | Temporal,<br>N = 8<sup>1</sup> | Temporal/basal/<br>occipital,<br>N = 10<sup>1</sup> | Anterior<br>perisylvian,<br>N = 20<sup>1</sup> | Perisylvian,<br>N = 22<sup>1</sup> | p-value<sup>2</sup> |
| --- | --- | --- | --- | --- | --- | --- |
| Sex |  |  |  |  |  | 0.3 |
| M | 33 (55%) | 4 (50%) | 3 (30%) | 13 (65%) | 13 (59%) |  |
| F | 27 (45%) | 4 (50%) | 7 (70%) | 7 (35%) | 9 (41%) |  |
| Age (years) | 34 [28, 44] | 34 [23, 39] | 32 [28, 48] | 37 [29, 48] | 33 [26, 38] | 0.5 |
| MRI abnormality | 40 (67%) | 3 (38%) | 8 (80%) | 14 (70%) | 15 (68%) | 0.3 |
| Number of electrodes | 14 [13, 15] | 11 [9, 13] | 15 [14, 15] | 14 [13, 15] | 14 [14, 16] | **0.044** |
| SEEG laterality |  |  |  |  |  | 0.4 |
| Right | 15 (25%) | 4 (50%) | 1 (10%) | 6 (30%) | 4 (18%) |  |
| Left | 24 (40%) | 1 (13%) | 4 (40%) | 8 (40%) | 11 (50%) |  |
| Bilateral | 21 (35%) | 3 (38%) | 5 (50%) | 6 (30%) | 7 (32%) |  |
| Additional electrodes<br>required | 1 (1.7%) | 0 (0%) | 0 (0%) | 1 (5%) | 0 (0%) | 0.57 |
| Post SEEG intervention |  |  |  |  |  | 0.9 |
| None | 4 (6.7%) | 0 (0%) | 1 (10%) | 2 (10%) | 1 (4.5%) |  |
| Resection | 49 (82%) | 7 (88%) | 7 (70%) | 16 (80%) | 19 (86%) |  |
| Neuromodulation | 7 (12%) | 1 (13%) | 2 (20%) | 2 (10%) | 2 (9.1%) |  |
| Extent of resection<sup>3</sup> |  |  |  |  |  | 0.3 |
| Temporal | 34 (69%) | 7 (100%) | 4 (57%) | 10 (63%) | 13 (68%) |  |
| Extratemporal ±<br>temporal | 15 (31%) | 0 (0%) | 3 (43%) | 6 (38%) | 6 (32%) |  |
| Resection volume (cc)<sup>3</sup> | 30 [22, 40] | 40 [29, 41] | 31 [21, 35] | 28 [17, 38] | 26 [22, 32] | 0.4 |
| Seizure freedom<sup>3</sup> | 36 (73%) | 6 (86%) | 5 (71%) | 12 (75%) | 13 (68%) | 0.9 |

<sup>1</sup>n (%); Median (IQR)<sup>2</sup>Pearson's Chi-squared test; Kruskal-Wallis rank sum test; Fisher's exact test<sup>3</sup>Seizure freedom, resection volume computed over resection patients only

### Classification of TLE hypotheses and SEEG explorations

We proposed a classification system for hypotheses and subsequent patterns of SEEG explorations in TLE consisting of 4 types - 1) Mesial/Lateral Temporal (MLT), 2) Temporal/Basal/Occipital (TBO), 3) Anterior Perisylvian (AP), and 4) Perisylvian (P). Both the cytoarchitectonic parcellation of the temporal lobe and each patient’s anatomical, semiological, and non-invasive electrophysiological data were considered.

Each hypothesis leads to a characteristic SEEG implantation. Multiple hypotheses, as consequence, will result in combinations of different patterns of implantation. Each classification is described further in detail. Anatomic illustration and case examples are shown in **Figure 1**.

**Figure 1:**
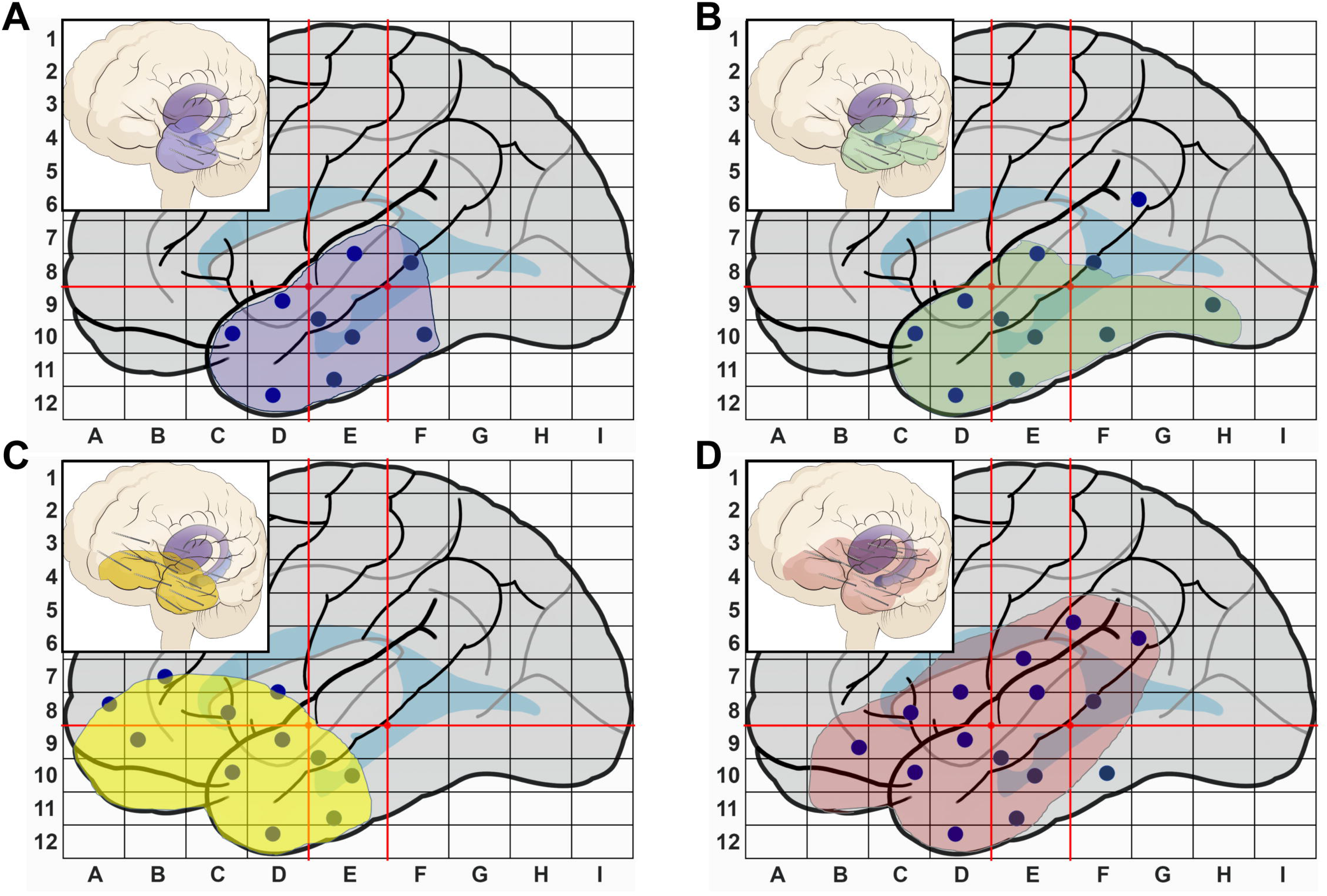
**Classification of temporal lobe epilepsy SEEG implantation hypotheses**. Talairach grids demonstrating implantation schemes in an example patient for each classification. *Inset:* 3D representation of an idealized left-sided implantation for each hypothesis demonstrating orthogonal lateral-to-medial trajectories and interrogation of cortical and subcortical structures. A: *Mesial/lateral temporal*: Epileptogenic zone is hypothesized to be in the mesial and/or lateral compartments of the temporal lobe with implantation focused on the hippocampal formation, entorhinal cortex, perirhinal cortex, amygdala, temporal pole, and lateral temporal neocortex. B: *Temporal/basal/occipital*: This hypothesis anatomically involves the posterior and basal temporal regions with extension to the occipital lobe. In addition to temporal coverage, the implantation covers posterior inferior temporal gyrus, fusiform gyrus, occipital lobe, temporo-parietal- occipital (TPO) junction, retrosplenial cortex, and posterior cingulate cortex. C: *Anterior perisylvian*: The epileptogenic zone involves the paralimbic regions including anterior temporal lobe, rostral insula and orbitofrontal regions and the SEEG covers these regions. D: *Perisylvian*: These hypotheses are characterized by the involvement of the anterior and posterior perisylvian regions, the TPO junction, and parietal lobes. SEEG coverage extends through the temporal lobe, parietal lobe, TPO junction, and frontal and occipital regions as necessary.

Additional case examples with semiological and surgical details can be found in Supplementary Results (**Supplementary** Figure 4 and 5).

I). Mesial/lateral temporal (MLT): The MLT hypothesis (**Figure 1A**) is proposed when the pre- implantation workup suggested that the EZ zone was confined in the mesial/lateral compartments of the temporal lobe. Semiologically, it is non-restrictively characterized by minimal ictal manifestations, and potentially by autobiographic (déjà vu or dreamy state), gustatory or autonomic auras. Here, the SEEG implantation is focused on the hippocampal formation, entorhinal cortex (Brodmann Area 34, 28, 27), perirhinal cortex (BA 35, 36), amygdaloid complex, the temporal pole (BA 38) and the lateral temporal neocortex as the anterior aspects of the superior, middle, and inferior temporal gyri (BA 22, 21, 20, respectively), usually no further posteriorly than Heschl’s gyrus (BA 41, 42). As electrodes are implanted orthogonally, both mesial and lateral temporal structures are explored concomitantly. Typically, a median of 11 [9,13] electrodes were utilized to interrogate this hypothesis. Implantation patterns exploring other hypothesis types in the temporal lobe and adjacent regions generally build upon the MLT pattern.
II). Temporal/basal/occipital (TBO): TBO hypotheses (**Figure 1B**) can be broadly characterized by involvement of posterior and basal temporal lobe regions with extension to the occipital lobe. Non-invasive data suggests the involvement of the temporal lobe, but with additional semiological, electrophysiological and/or imaging data suggesting the possible involvement of the basal and dorsal-lateral temporal/occipital areas. Commonly, these additional ictal signs and symptoms were associated with the presence of visual auras, conjugate eye deviation, and, if on the dominant side, language disruption. This approach extends the MLT implantation to the basal temporal lobe and fusiform gyrus (BA 37), occipital lobe (BA 17-19), temporal-parietal-occipital (TPO) junction (BA 39, 40), retrosplenial cortex (BA 29, 30), and posterior cingulate cortex (BA 23, 31). The temporal/basal/occipital explorations were achieved with 15 [14,15] electrodes implanted orthogonally.
III). Anterior perisylvian (AP): The AP hypothesis (**Figure 1C**) interrogates the involvement of the paralimbic regions in the organization of temporal lobe seizures. Typically, the exploration includes the anterior temporal lobe (including the mesial temporal structures as the amygdala and the anterior hippocampus), rostral ventral aspects of the insula (BA 13), orbitofrontal regions (BA 11, 12), frontal pole (BA 10), inferior frontal gyrus (BA 44, 45, 47), and subgenual cingulate cortex (BA 25). If more prominent naturalistic and integrative motor manifestations are prominent, additional frontal lobe structures including premotor (BA 6, 8), anterior cingulate (BA 32, 33) and primary motor (BA 4) are added. These cortical areas are structurally interconnected via the uncinate fasciculus, surrounding the sphenoidal surface of the Sylvian fissure, surrounding the *limen insulae*. Semiological features can include fearful behavior, oral automatisms, autonomic symptoms, and other limbic manifestations. The AP explorations were achieved with median 14 [13, 15] orthogonal electrodes.
IV). Perisylvian (P): Perisylvian hypotheses (**Figure 1D**) were characterized by the contribution of anterior and posterior perisylvian regions, possibly involving the dorsal-lateral temporo- parietal-occipital (TPO) cortex and vicinity. Due to the multimodal associative characteristics of the cortical organization in this region, semiological features are complex and many times difficult to interpret. Manifestations can include dizziness, conjugate eye deviation, extremities sensory symptoms, sensory and motor face and throat manifestations, auditory phenomena and speech disturbance. SEEG sampling includes the TPO junction and associated temporal and parietal operculum (BA 39, 40), superior parietal lobule (BA 5, 7), perisylvian primary sensory cortex (BA 3, 1, 2), retrosplenial cortex (BA 29, 30), posterior cingulate cortex (BA 23, 31), perisylvian primary motor cortex (BA 4), and inferior frontal gyrus (BA 44, 45). A median of 14 [14, 16] electrodes were used for this exploration.

### Quantitative validation of semiological features and hypothesis classification

The frequencies of semiological features by hypothesis classification are reported in **Figure 2** and semiology by individual patients in **Supplementary** Figure 6. Using RFE, the 10 most predictive features were selected (**Figure 2, Supplementary Table 2**). Our quantitative approach identified several features consistent with our conceptualization of the proposed classification.

**Figure 2:**
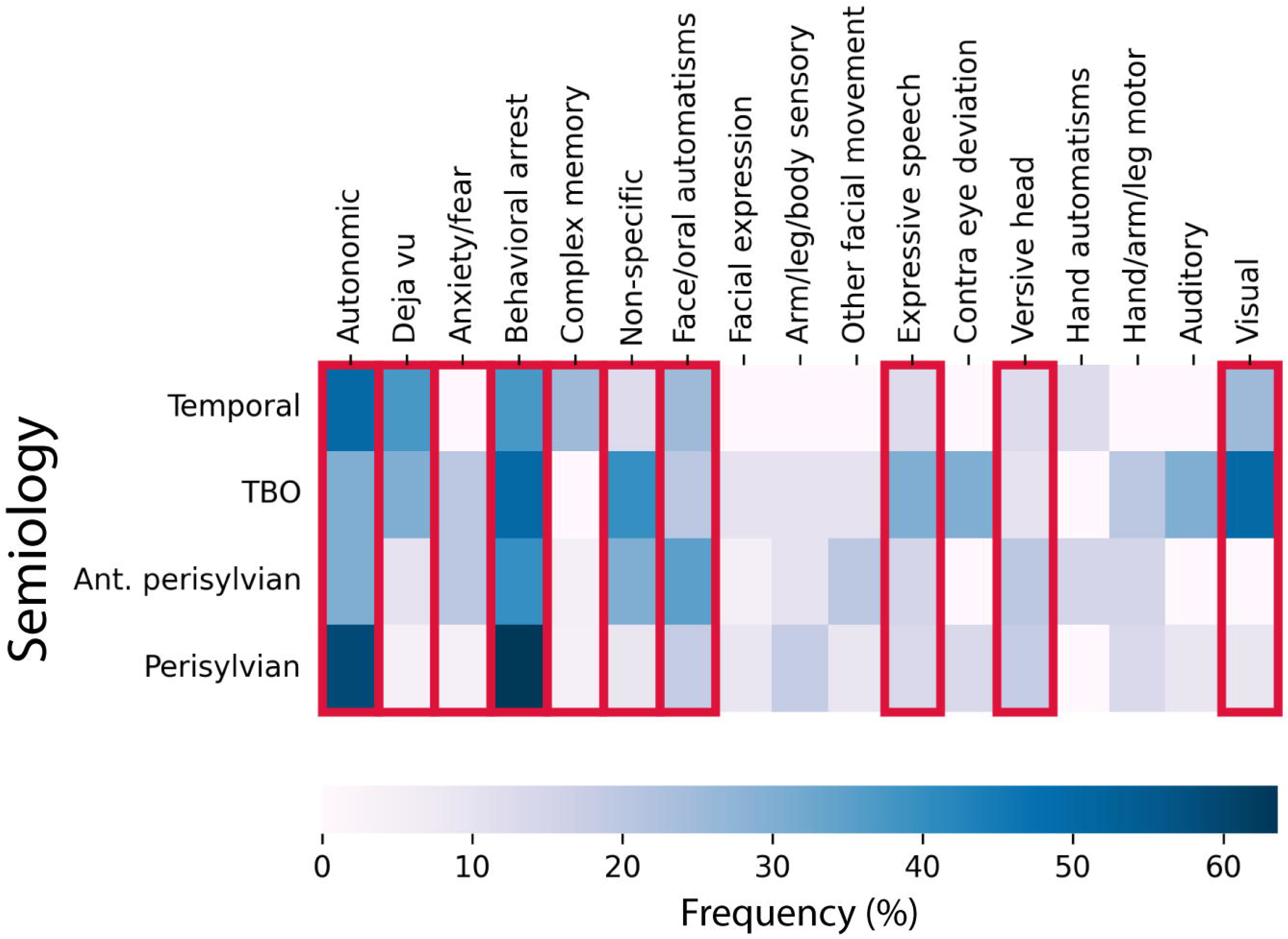
**Semiology frequencies by classification**. Red outline delineates the ten most features that had the most predictive power in differentiating classifications by recursive feature elimination (RFE). We quantitatively validated classification schemes by determining the overall accuracy of random forest classification with the features determined by RFE analysis. The classification was developed using AEC features of various patients with temporal lobe epilepsy. RFE analysis demonstrates accordance with qualitatively differentiating features (for instance visual phenomena in temporal/basal/occipital or facial/oral automatisms in anterior perisylvian). Random forest classification (500 trees, Gini impurity criterion) achieved 90% classification accuracy on the dataset of 60 patients. Individual patterns are reported in **Supplementary** Figure 6.

Déjà vu (33%) and dreamy state (50%) were most common in MLT. TBO was associated with visual symptoms (56%) and expressive speech disturbances (30%). AP had high proportion of facial/oral automatisms (47%), versive head movements (40%), non-localized/generalized sensation (46%) and anxiety/fear (56%). Finally, P was associated with behavioral arrest (47%), autonomic changes (50%), versive head movements (40%), and arm/leg/body lateralized sensory changes (23%). We used the 10 selected features to predict the hypothesis classification of each subject, based on semiology alone. A random forest classifier classified the cohort with 90% accuracy.

### Electrode locations in stereotaxic space

We collected the Talairach and MNI coordinates of the most distal contact of each electrode (**Supplementary Table 1**). There was a high degree of consistency between implantations that corresponded with our AEC hypotheses (**Figure 3A**). Coronal cross sections showing typical trajectories is shown in **Supplementary** Figure 3.

**Figure 3:**
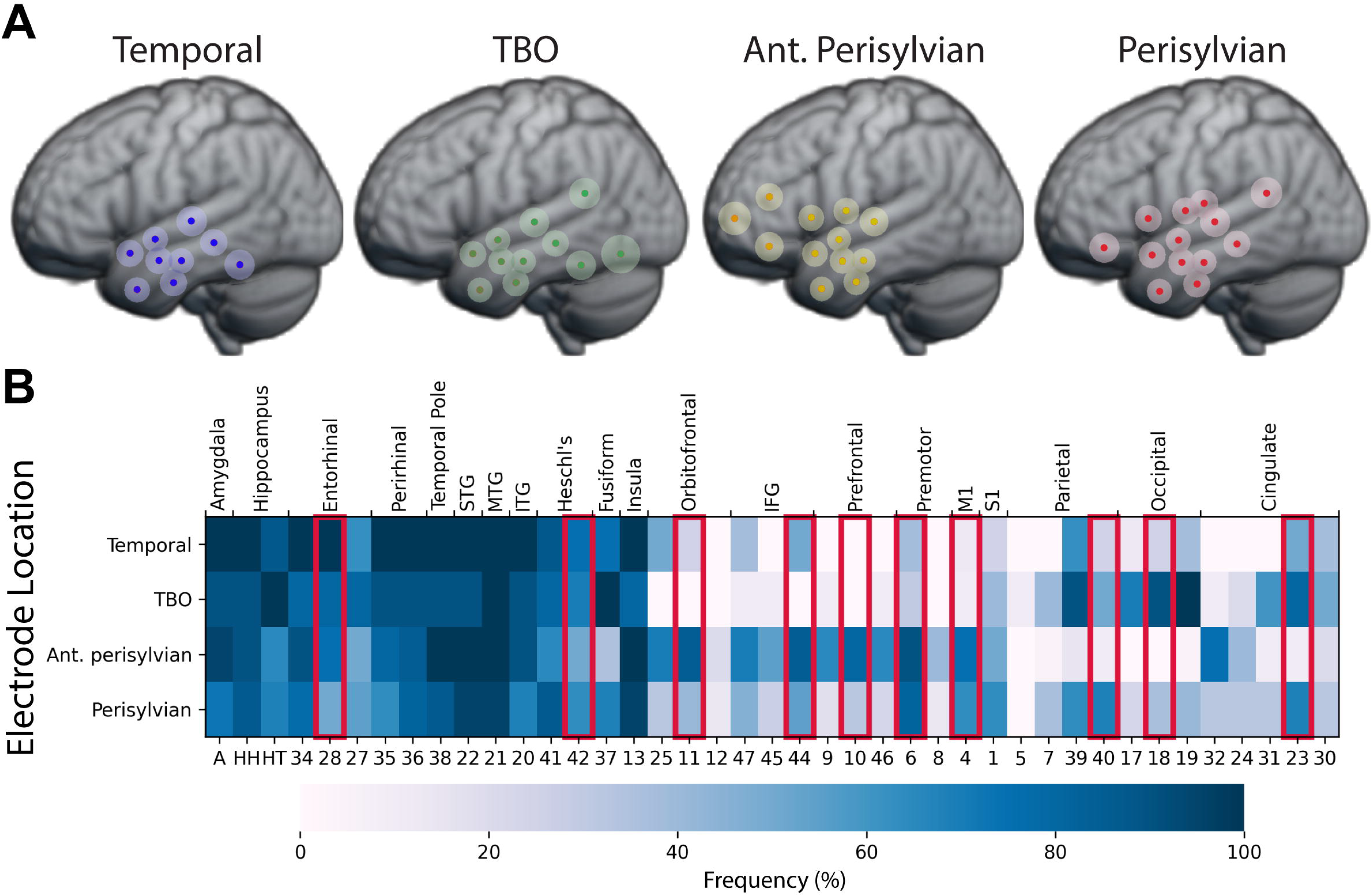
**Electrode locations**. Electrode locations were extracted from post SEEG placement CT scans and registered to preoperative patient MRI and then to the MNI atlas. We calculated the Talairach and MNI coordinates of the medial most contact in each electrode for each patient. As electrodes are implanted orthogonally to the dorsolateral cranial surface, this fixes the position of the electrode, with minor changes in entry point to prevent vascular collisions. Brodmann areas were extracted in a semi-automated fashion as described in the methods. Coordinates are described quantitatively in **Supplementary Table 1**. A. *Bubble plots* of median (dot) and IQR (bubble) of target contact coordinates (MNI), displayed over the MN152 brain surface. Note the high degree of consistency between electrode targets. B. *RFE analysis* successfully identified features predictive of each classification. These were expectedly largely extratemporal locations and corresponded to the anatomic localization of each hypothesis, e.g. frontal regions for anterior perisylvian, occipital regions for temporal/basal/occipital, etc. Random forest classification (500 trees, Gini impurity criterion) achieved 97% classification accuracy. *Bottom x-axis – Brodmann regions, Top x-axis – areas grouped by region.* Individual patterns are reported in Supplementary Figure 6.

Electrode coverage is shown in aggregate in **Figure 3B** and in individual subjects in **Supplementary** Figure 7. Using RFE, we identified the 10 electrode locations with the most predictive ability to distinguish classes (**Figure 3B**, **Supplementary Table 2**). As the electrodes within the temporal lobe were similar between classifications, most selected electrodes were extratemporal. Specifically, occipital (90%) and posterior cingulate (80%) were most common in TBO. Prefrontal (80%), inferior frontal gyrus (85%), premotor (90%), primary motor (75%), and orbitofrontal (85%) were most common in AP. TPO/parietal (68%), posterior cingulate (68%), premotor (82%), and primary motor (64%) electrodes were most common in P. The random forests classifier achieved 97% accuracy in determining the hypothesis based on electrode location.

We evaluated the frequency of each classification at combinations of semiology-electrode features (**Figure 4**). This demonstrated strong visual clustering, consistent with our previous findings of cardinal semiological and electrode location features for each hypothesis classification.

**Figure 4:**
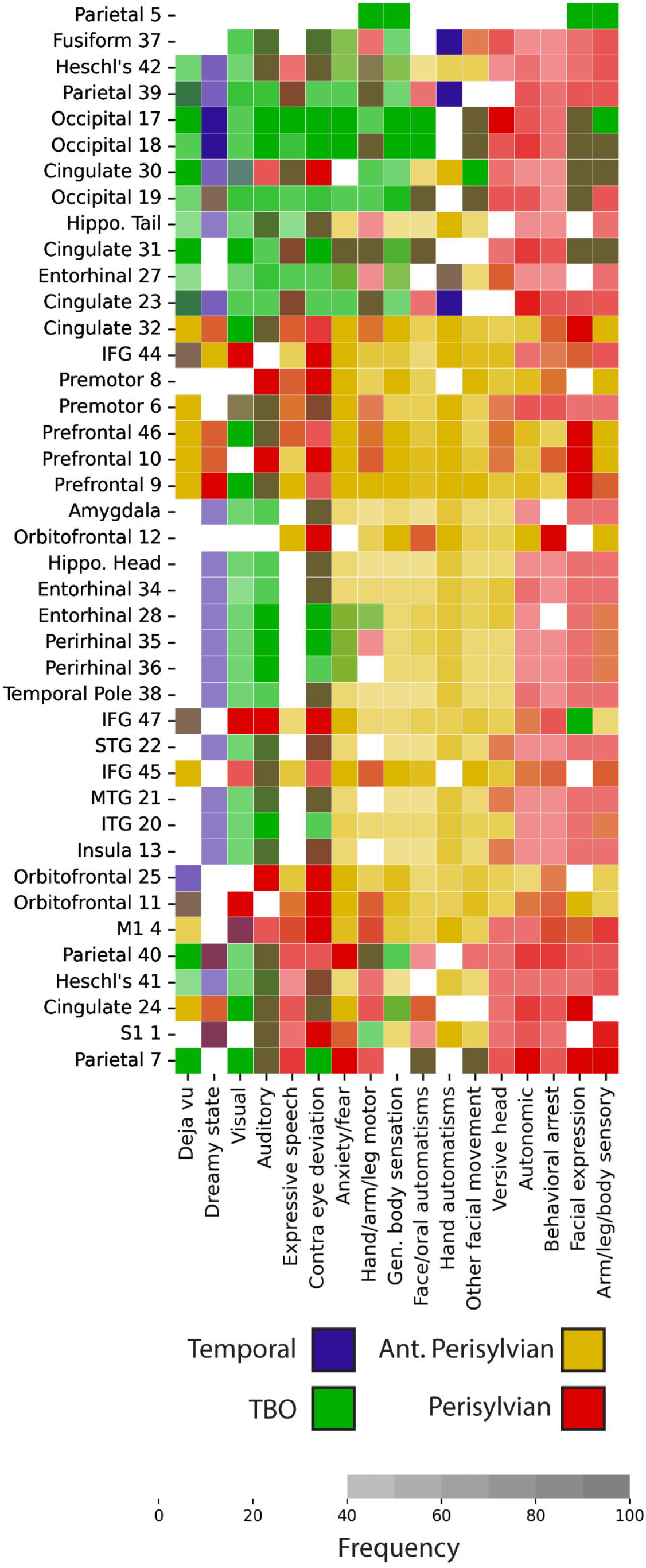
**Frequency of semiology-location pairs by classification**. The frequency of classification by pairs is demonstrated and shows strong clustering. This clustering largely corresponds to the prior RFE/random forest classification. Most pairs were dominated by one classification with mixed colors indicating that 2 classifications had equal frequencies. Blank cells indicate that no classification had greater than 40% frequency.

### Post SEEG Intervention

Following SEEG, 82% underwent surgical resection, 12% underwent neuromodulation, and 6% did not undergo additional intervention. 69% of resection patients underwent tailored temporal resections, while 31% underwent extratemporal with or without temporal resections. Individual resection patterns are illustrated in **Figure 5**. The median resection volume was 30 [22, 40]cc and did not vary significantly between classifications (p=0·4), nor did the degree of extratemporal vs. temporal resection (**Table 1**; p=0·3). No MLT implantations resulted in extratemporal resections. Very few patients underwent a “standard temporal lobectomy” (resection of all temporal structures to 4·5/5·5 cm from the temporal tip) in our series. For instance, of 49 patients, 29 (59%) had the hippocampal head resected and12 (24%) had the hippocampal body/tail resected. Only 3/60 patients had resections that could be characterized as standard temporal lobectomies. Using the RFE feature selection approach to areas of resection (**Supplementary Table 3**), the overall classification accuracy using a random forest classifier was 63%

**Figure 5:**
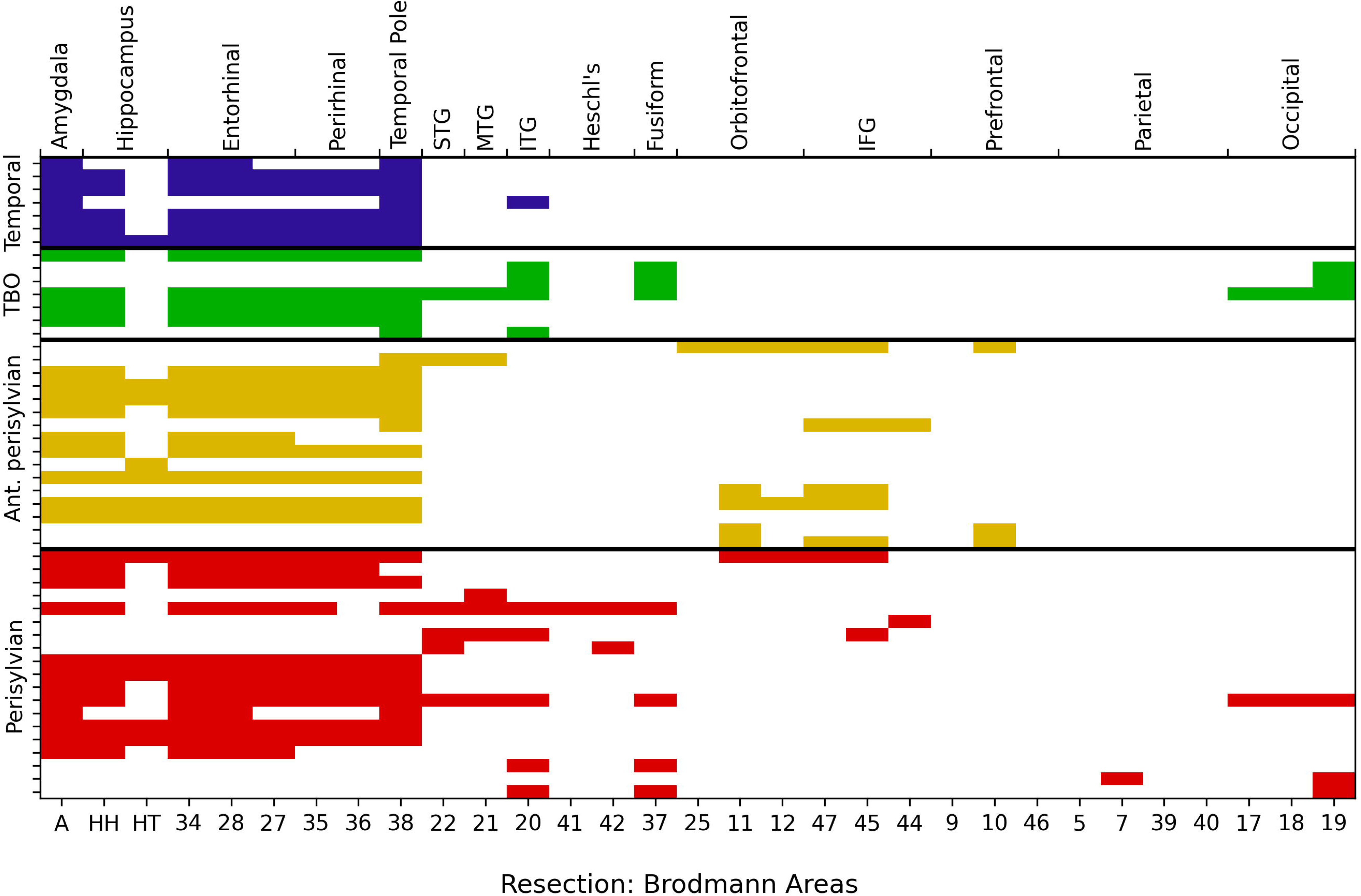
Resection patterns by classification. 49 of 60 patients underwent resection with an overall one-year seizure freedom rate of 73% (temporal: 86%, TBO: 71%, anterior perisylvian: 75%, perisylvian 68%, p=0.9). Each row represents an individual patient while filled cells indicated resected regions, colored by classification as in Figure 1. *Temporal* (N=7): All patients had resections entirely confined to the temporal lobe and largely had preservation of posterior temporal regions and the hippocampal tail. *Temporal/basal/occipital* (N=7): 5 patients (57%) resections confined to the temporal lobe, including posterior regions such as fusiform gyrus (BA37) and ITG (BA20) with 3 (43%) undergoing resections of occipital with or without temporal lobe regions. *Anterior perisylvian* (N=16): 10 (63%) patients underwent resection of largely anterior temporal regions while 6 (36%) underwent resection of additional inferior frontal lobe structures. *Perisylvian* (N=19): 13 (68%) underwent resection of temporal regions, including posterior regions, while 6 (32%) underwent additional resection of frontal/parietal/occipital regions.

### Seizure freedom

Of those who underwent resection, the one-year seizure freedom (Engel 1) rate was 73% and did not vary by classification (MLT: 86%, TBO: 71%, AP: 75%, P: 68%; p=0.9), sex, age, MRI abnormality, number of electrodes, or laterality of implantation (**Supplementary Table 4**). One-year seizure freedom was significantly associated with post SEEG intervention (100% resection in seizure free vs. 65%/35% resection/neuromodulation in not seizure free, p<0.001).

### Relationship of MRI lesions to implantation patterns

MRI abnormalities suggestive of hippocampal sclerosis, focal cortical dysplasia, among others were found in 67% of patients and did not vary by classification (p=0·3, **Table 1**). The patterns of implantation are primarily informed by the AEC hypotheses of localization as described before. The presence of an MRI visible lesion does play a role in surgical planning, but lesions minimally influence the placement and trajectory of SEEG electrodes. MRI lesions did not affect seizure freedom (not seizure free - 60% vs. seizure free - 69%, p=0·5, **Supplementary Table 4**).

### Neuropsychological outcomes

Neuropsychological testing was obtained pre-operatively in all patients as part of Phase I evaluation and for all resection/ablation patients at 6-months post-resection, with variable follow-up (25/49 resection patients). Decline/improvement is defined as change >1 S.D. from preoperative performance. Verbal memory declined in 26% of patients (36% left, 11% right), and improved in 13% (14% left, 11% right). Picture naming declined in 19% (29% left, 0% right), and improved in 9·5% overall (0% left, 29% right). Finally, visuospatial memory declined in 8% of patients (13% left, 0% right), and improved in 24% (19% left, 33% right) (**Figure 6**). Other measures, including reading and semantic/phonemic fluency are reported in **Supplementary Table 5**.

**Figure 6:**
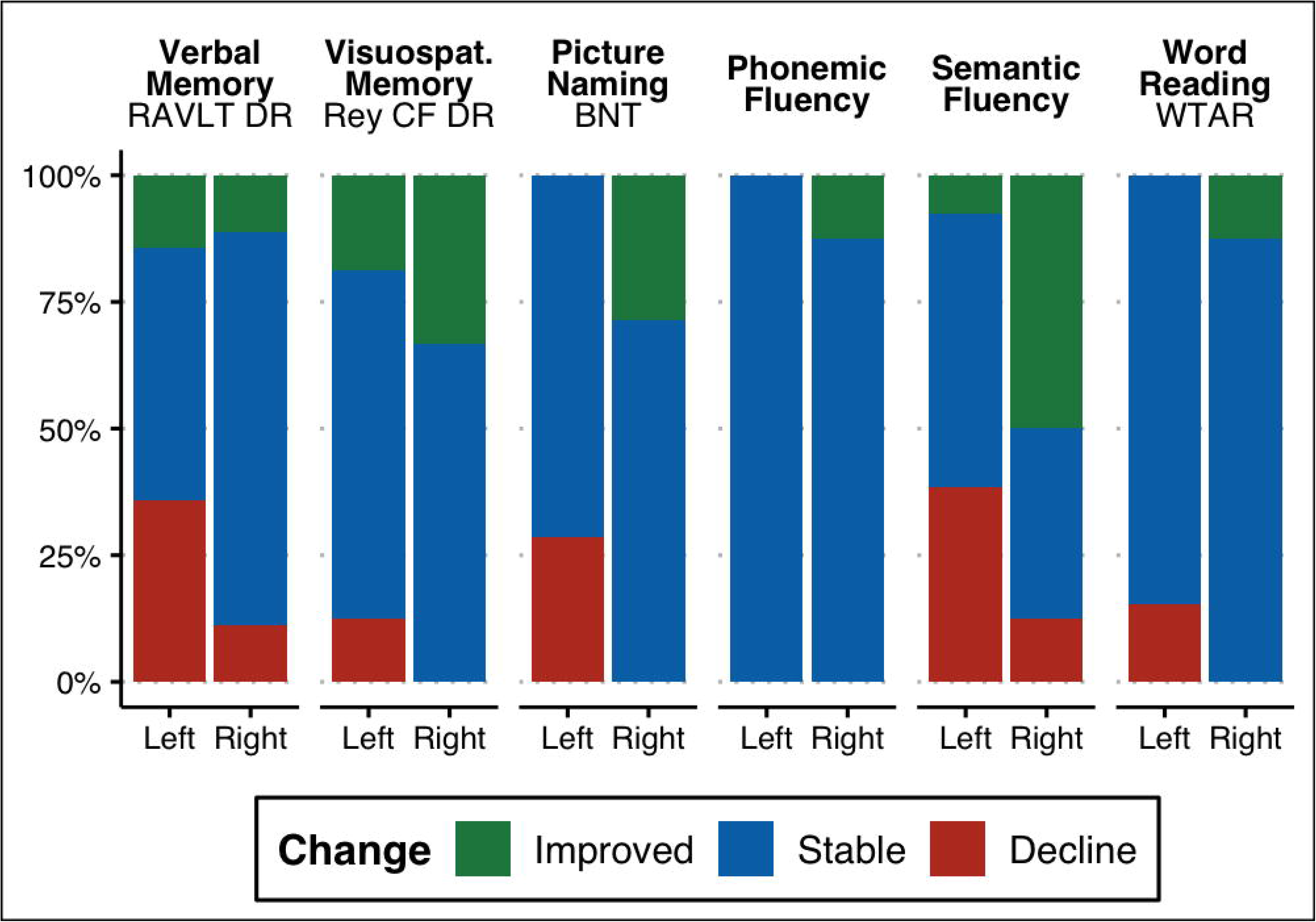
Neuropsychological outcomes. Six-month neuropsychological outcomes in patients who underwent surgical resection or ablation of the EZ by side of intervention. Verbal memory (RAVLT delayed recall) declined in 26% of patients overall (36% left, 11% right), and improved in 13% (14% left, 11% right). Visuospatial memory (Rey-Osterrieth Complex Figure delayed recall) declined in 8% of patients (13% left, 0% right), and improved in 24% (19% left, 33% right). Picture naming (Boston Naming Test/NAB Naming Test) declined in 19% of patients (29% left, 0% right), and improved in 9.5% (0% left, 29% right). Phonemic fluency declined in no patients. Semantic fluency declined in 29% of patients (38% left, 13% right). Word reading (Wechsler Test of Adult Reading) declined in 9.5% (15% left, 0% right)

## Discussion

Here, we develop, qualitatively describe, and quantitatively validate a proposal for classifying hypotheses in TLE. Our approach is predicated upon the integration of specific seizure semiological features and anatomo-clinical correlations, with meticulous semiological analyses serving as the cornerstone in this classification and implantation design process. At the heart of our approach lies an effort to introduce reproducible, orthogonally oriented trajectories, with strict adherence to the foundational principles of SEEG methodology. It is imperative to underscore the inherent variability in patients with TLE, particularly in the nuanced semiological aspects. Hence, while our proposed hypotheses concerning localization and subsequent patterns of implantation serve as initial frameworks, they necessitate supplementation, amalgamation, and modification to accommodate individual idiosyncrasies gleaned from the pre-implantation data. The presence of an MRI lesion minimally modified the implantation strategy, possibly with the addition of one or two trajectories to explore the role of the lesion in the organization of the epileptic activity.

Despite the inherent challenges associated with standardizing the management of such individualized conditions as TLE, our study underscores the pivotal role of fundamental semiological features and clear hypotheses guiding the precise placement of SEEG. Our approach aims to bridge personalized medicine and the imperative for standardized approaches to optimize outcomes in epilepsy surgery. Only one of 60 patients required a return to the operating room for placement of additional electrodes, highlighting the appropriate coverage of the original hypotheses. Subsequently, surgical resection led to a 73% overall seizure freedom rate at 1 year with only 26% verbal memory decline and improvement in 13%, using a selective, patient-tailored surgical approach. To achieve a practical realization of this approach, we provide standardized targeting coordinates for each electrode trajectory.

Resection target did not predict hypothesis classification well (63% accuracy) in comparison to prediction with semiological features or electrode location. This is concordant with the purpose of SEEG and validate the importance of SEEG explorations in TLE when non-invasive data does provide a well-defined surgical strategy. If hypothesis classification based on non-invasive data could predict the location and extent of EZ (e.g. resection target) with high accuracy, there would be no need for intracranial exploration, and the patient could proceed directly to surgical resection. In the SEEG methodology, one or more AEC hypotheses are proposed and specifically interrogated, and the ultimate surgical plan (if appropriate) is then determined based on this data. This may also explain the low rate of mesial/lateral temporal hypotheses in our study cohort (8/60, 13.3%). These patients often have highly characteristic semiology and concordant imaging and electrophysiological findings and proceed direct to surgical interventions without intracranial evaluation.

SEEG explorations using this framework permitted accurate delineation of the location and extent of surgical resections. Most patients did not undergo a “standard temporal lobectomy”, with only a small fraction (3 patients out of 60) undergoing such procedures. Despite this focused approach, seizure freedom remained remarkably favorable (73% overall, 86% in the MLT group). Notably, these resections predominantly encompassed subregions of the temporal lobe, and extended into adjacent cortical regions in 31% of cases, including the insula or segments of the frontal, parietal, or occipital lobes. This finding underscores a fundamental paradigm shift: temporal lobe epilepsy transcends mere anatomical localization, evolving into a complex functional-anatomical construct necessitating tailored surgical strategies over a standardized approach.

Our surgical approach led to improved neuropsychological outcomes compared to other reports. For instance, verbal memory decline occurred in 36% of patients after left sided surgery and 11% after right surgery in this cohort, compared to reported rates after temporal lobectomy of 44% on the left and 11% on the right^20^. In fact, the verbal memory decline rates are comparable to those after laser ablation in a recent large series (35·3% left, 16·3% right)^21^. The neuropsychological consequences of selective approaches is a controversial topic with some reports finding benefit from selective (largely mesial) approaches over standard temporal lobectomies and other showing no significant difference^22–25^. Somewhat in contrast, in one report, sparing of the posterior hippocampus was associated with significantly reduced odds of postoperative verbal memory decline^26^. Here, our targeted approach spared the posterior hippocampus in 76% of cases, permitting this potential benefit. That said, seizure freedom and improvement in quality of life are the ultimate aims and *appropriately* selective interventions should maximize seizure freedom while minimizing neuropsychological morbidity.

The implantation patterns are further concordant with the white matter anatomy of the temporal lobe and surrounding structures as well as anterior-posterior gradients in hippocampal/parahippocampal connectivity. The TBO pattern mirrors the course of the inferior longitudinal fasciculus and inferior frontal occipital fasciculus. Anterior perisylvian implantations explore paralimbic regions connected by the uncinate fasciculus, while perisylvian implantations follow the course of the arcuate fasciculus. AP and P classifications build on prior descriptions of “temporal perisylvian” epilepsies^27^ , but are divided by the presence of specific semiological features (as described previously) and posterior extension into the posterior insula, parietal lobes, and/or TPO junction. The distinction between AP and TBO/P patterns is also supported by the extensive literature contrasting anterior and posterior hippocampal/parahippocampal regions^28–30^. Thus, our classification may represent functionally distinct pathological networks in TLE, worthy of further focused structural and electrophysiological research.

The proposed classification of hypothesis-driven patterns for SEEG implantation in TLE synthesizes the authors’ clinical expertise from over 850 SEEG implantations over 15 years and is validated with detailed semiological and structural analyses of a cohort of 60 patients. The classification is validated using a quantitative, machine learning-based approach. Our recommendation is that this classification system serves as a starting point for AEC correlations in individual patients, rather than a definitive or restrictive guide. Tailoring the SEEG explorations for each patient by integrating multimodal information and multidisciplinary discussion will result in more selective and volumetrically-restricted surgical interventions that maximize seizure freedom while minimizing neuropsychological morbidity.

## Funding

This work was funded partially by NIH 1F32DC020644-1 to ANM, NIH 1R01NS122927-01A1 to JGM, and funding via the Physician-Scientist Institutional Award from the Burroughs Wellcome Fund to the University of Pittsburgh. Funding sources were not involved in the data collection, analysis, writing of the manuscript, or decision to submit it for publication. No work was done for hire from a pharmaceutical company or other agency. Authors were not precluded from accessing data in the study and all authors accept responsibility to submit for publication.

## Declaration of interests

JGM is a consultant for Zimmer Biomet. The authors report no other interests.

## Data sharing statement

Data collected for this study, including deidentified clinical data, electrode coordinates, seizure outcomes, semiological scoring, resection locations, and code are available upon reasonable request to the corresponding author.

## Author contributions

ANM, TA, JGM conceived the study. ANM, JH, ER, NI, HA, TC, and AA performed data collection. LCH and DRC performed neuropsychological testing and analysis. ANM, JH, ER, NI, and TA performed data analysis. ANM, JH composed the manuscript. ANM, JH, ER, NI, HA, and TC assembled figures and tables. ANM, JGM, TA, and HA critically revised the manuscript. JGM supervised the study. All authors met ICMJE criteria for authorship and approved the manuscript in its final form.

## Data Availability

All data produced in the present study are available upon reasonable request to the authors

## Notes

### Author Declarations

The IRB of University of Pittsburgh Medical Center gave ethical approval for this work (STUDY #21020058).

