## Supplementary Material for "Hypothesis-driven patterns of intracranial exploration in temporal lobe epilepsies"


**Supplementary Methods** 2
Supplementary Figure 1 3
Supplementary Figure 2 4
**Supplementary Results** 5
Supplementary Figure 3 5
Supplementary Figure 4 7
Supplementary Figure 5 10
Supplementary Figure 6 12
Supplementary Figure 7 13
Supplementary Table 1 14
Supplementary Table 2 15
Supplementary Table 3 16
Supplementary Table 4 17
Supplementary Table 5 18
**References** 19

**Supplementary Methods**

*Phase I evaluation*

All patients were referred for Phase I non-invasive evaluation by the treating epileptologist. Phase I evaluation included inpatient scalp video EEG monitoring, high-resolution 3T structural MRI, magnetoencephalography, interictal PET, ictal/interictal SPECT, and neuropsychological evaluation. This information was then reviewed and discussed at our institution's multidisciplinary epilepsy patient management conference, which included epileptologists, neurosurgeons, neuroradiologists and neuropsychologists. The hypotheses for the EZ location and its extent, and subsequent decision for intervention (invasive Phase II monitoring, surgical intervention, or continued medical management) were developed by consensus at the conference.

If the patient was referred for Phase II SEEG monitoring, the SEEG trajectory planning was also determined by the conference based on working hypotheses. The working hypotheses guiding the SEEG trajectories took into consideration the conceivable anatomical EZ location and its extent, MRI lesions that were likely related to the organization of seizures, and possible anatomical location of relevant functional cortical areas.

*Neuropsychological Testing*

All patients underwent preoperative neuropsychological evaluation by a board eligible or board certified neuropsychologist (L.H., D.C.) including tests of verbal memory (Rey Auditory Verbal Learning Test Delayed Recall – RAVLT DR), visuospatial memory (Rey–Osterrieth Complex Figure Delayed Recall – ROCF DR), picture naming (Boston Naming Test – BNT or Neuropsychological Assessment Battery Naming Test – NAB), phonemic fluency, semantic fluency, and word reading (Wechsler Test of Adult Reading). Patients who subsequently underwent resection/ablation of the EZ were scheduled for 6-month postoperative (7 [6-9] month actual follow-up) neuropsychological testing of similar domains. A decline/improvement was defined as greater than 1 S.D. decrease/increase in the standardized score for each test. Patients with missing neuropsychological follow-up were excluded from analysis of neuropsychological outcomes.

*Preoperative planning and surgical technique*

After the working hypotheses were defined, the intended trajectories of electrodes were planned. Using the Talairach stereotaxic space as the primary framework, the trajectories intended to explore the cortical and subcortical structures in a three-dimensional conceptualization considered the distal, intermediate, and proximal cortical subcortical structures traversed by the intracranial electrode. Subsequently, the SEEG intended trajectories were planned on the ROSA system (Zimmer Biomet, Warsaw, IN) using preoperative high-resolution CT, MRI T1 with and without contrast, and CT angiography. Trajectories were planned to maximize gray matter coverage and avoid vascular collisions. Consistent with prior reports[1,2], the majority of electrodes were placed orthogonally. Small adjustments (particularly at the entry point) were made to avoid vascular, particularly venous, collisions. Preoperative images were registered and reoriented along the AC-PC line to facilitate planning in Talairach stereotaxic space.

Electrode implantation was performed as previously described[3,4]. The patients were placed in a Leksell G frame and attached to the ROSA robot. After laser-based registration and verification of registration accuracy, skin incision and skull drilling were performed with a Stryker drill (Strkyer, Kalamazoo, MI) with 2·5mm bit through the close-fit ROSA adaptor. The metal guiding fixation devices (Dixi Medical, Marchaux-Chaudefontaine, France) were securely screwed into the skull following which the metal stylets were passed to the planned depth. Subsequently, 0·8mm diameter DIXI electrode with the desired number of electrode contacts is placed. Following the SEEG procedures, high resolution (0·625mm isotropic resolution) CTs were obtained to assess electrode contact locations and for postoperative hemorrhages. After EMU monitoring phase, electrodes were removed, and the patient was discharged. The decision for subsequent interventions were again made by the multidisciplinary epilepsy patient management conference.

*Electrode trajectories*

All hypothesis classifications usually entailed exploration of the hippocampal formation (anterior and posterior regions), amygdala, entorhinal cortex and temporal pole areas, concomitantly with the exploration of the correspondent lateral and basal temporal cortical areas using orthogonal trajectories. These 5 orthogonal trajectories, which are anatomically described as follows:

1. **Temporal Pole**: Interrogation of Brodmann Area 38 is achieved by the implantation of 1 or 2 orthogonally placed electrodes located anterior to the vertical AC line, in the D12 or C10 Talairach coordinate system. Here the trajectories will explore the mesial and lateral aspects of the temporal pole region. Frequently, the C10 trajectory also explore the posterior orbito-frontal

cortex (BA 25) using a paralimbic trajectory[1]

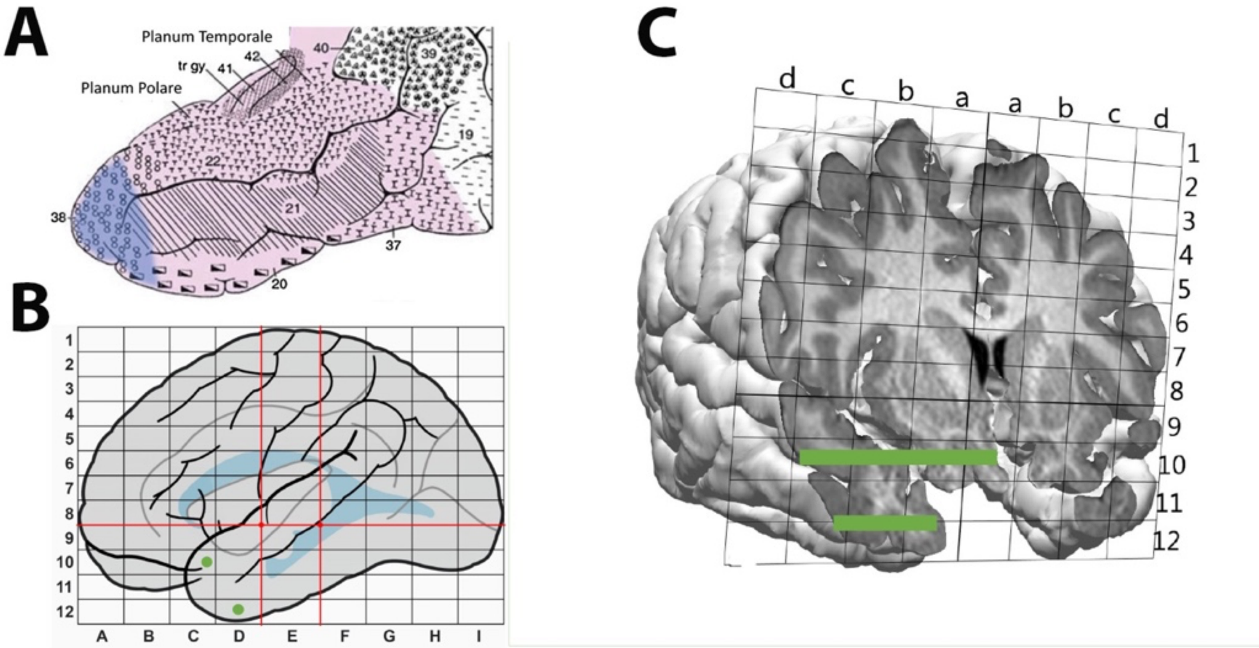


**Supplementary Figure 1: Temporal pole SEEG trajectories** – The inferior trajectory explores the basal/inferior aspects of the temporal pole while the super trajectory explores the superior temporal pole, paralimbic regions, and orbitofrontal cortices. A: Brodmann cytoarchitectonic map, B: Talairach grid representation, C: 3D reconstruction with superimposed Talairach grid

2. **Amygdala**: The amygdala is explored by electrodes implanted in orthogonal orientation, immediately posterior to VAC, at the most anterior areas of Talairach coordinate E10. At this coordinate, the trajectory will explore the anterior uncus and the basal lateral nucleus of the amygdaloid complex in the mesial aspect and the dorsal lateral neocortex (BA 21) in the lateral contacts. The amygdala is in proximity with VAC, approximately at 5mm posterior to the vertical line. The anterior uncus is located at the mesial projection of the amygdaloid nucleus, with the dorsal aspect corresponding to the piriform cortex. In the more lateral aspect of the orthogonal trajectory, the electrode explores the depths of the rostral aspect of the superior temporal sulcus and the crown of the middle temporal gyrus.
3. **Anterior Hippocampus**: This is achieved by electrodes implanted in orthogonal orientation, between the VAC and the VPC lines, in the E10 coordinate, but slightly posterior and ventral to the amygdala trajectory. At this coordinate, the trajectory will explore the posterior uncus and the head of the hippocampus in the most mesial aspect and BA 21 located in the rostral aspect of the middle temporal gyrus, in the most lateral aspect.
4. **Posterior Hippocampus**: To explore the posterior aspect of the hippocampus body and tail, the orthogonal trajectory is located at the F10 Talairach coordinate. This explores the posterior aspect of the parahippocampal gyrus, the tail of the hippocampus and the depth of the posterior aspect of the superior temporal sulcus, almost at the temporal/parietal/occipital transition, corresponding to the interface between Brodmann areas 22, 21, 39 and 19.
5. **Entorhinal cortex**: The entorhinal cortex, with its prominent connections with the head of the hippocampus and the amygdaloid nucleus are important structures to be explored in “dreamer states” scenarios. The same electrode that explores the entorhinal cortex on the mesial contact will cross the depths of the collateral sulcus in the more lateral locations, and subsequent explore the rostral fusiform gyrus, the depth of the temporal-occipital sulcus and will finally explore the inferior temporal gyrus, on tis most lateral trajectory. The trajectory is located immediately ventral to the hippocampus trajectory, at the E11 coordinate.

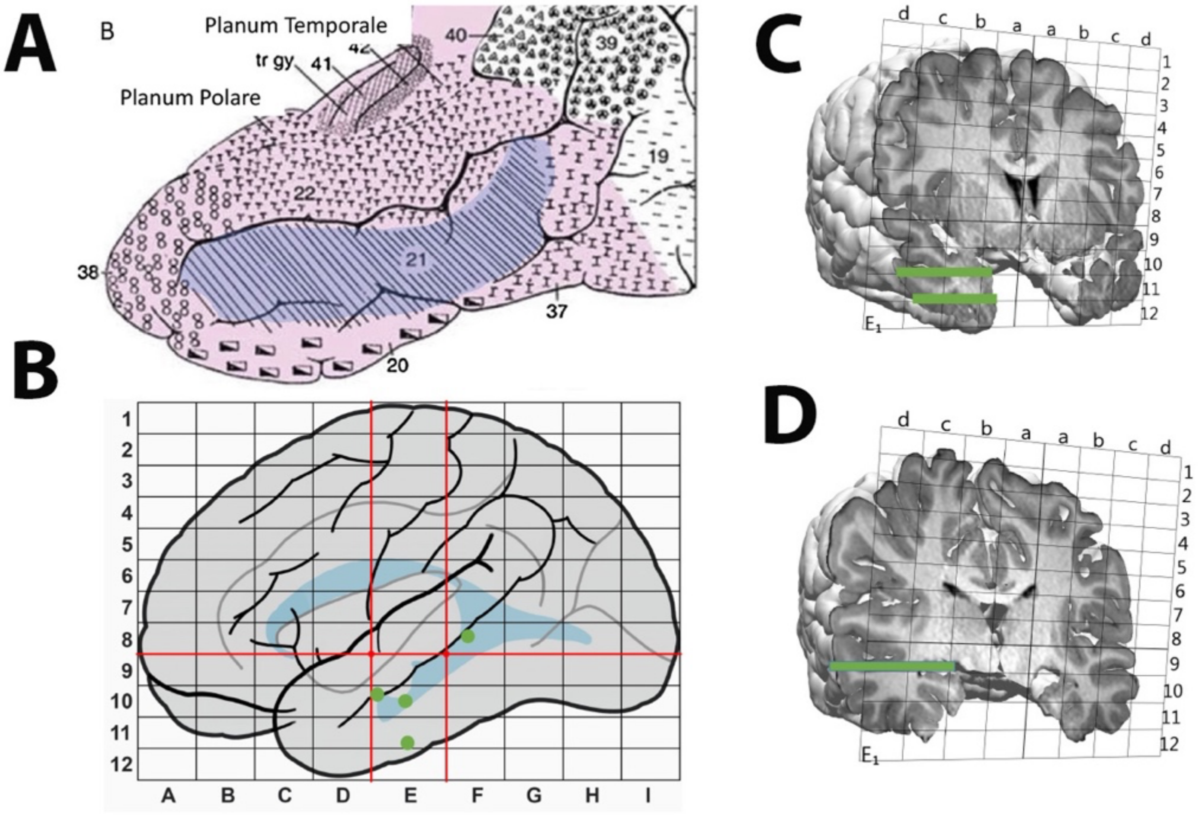


**Supplementary Figure 2: Amygdala, hippocampus, entorhinal cortex SEEG trajectories** A: Brodmann cytoarchitectonic map, B: Talairach grid representation, C: 3D reconstruction with superimposed Talairach grid of the amygdala and entorhinal cortex trajectories. D: 3D reconstruction of the hippocampal tail trajectory

*Electrode reconstruction and areas of resection*
SEEG electrodes reconstructions and coordinate extractions were performed using Curry (Compumedics Neuroscan, Charlotte, North Carolina, USA). Post-SEEG placement high-resolution CTs were registered to preoperative high-resolution MRIs and then to the MNI template. Reconstructions were reviewed by the surgical team to ensure appropriate localization of contacts. The Talairach stereotaxic coordinates of electrode contacts were converted to MNI coordinates and subsequently Brodmann areas using the label4MRI tool (https://github.com/yunshiuan/label4MRI/) which is built on the mni2tal tool in BioImageSuite (https://bioimagesuiteweb.github.io/bisweb-manual/tools/mni2tal.html) and is based off the registration in Lacadie 2009[5]. Brodmann areas were manually inspected for accuracy. For patients who underwent post-SEEG resection, postoperative high resolution T1 MRI was inspected against the Brodmann atlas by the surgical team and authors to determine which Brodmann areas were resected. Resection areas were manually segmented, and volume was calculated in the BrainLab (BrainLab AG, Munich, Germany). Electrode location and trajectory was illustrated over the MNI152 atlas[6].

*Statistical analysis and machine learning details*
Machine learning was performed using *scikit-learn*[7] in Python 3·10. Heatmaps and other figures were generated using *seaborn*[8]. Other statistical analyses were performed in R 4.2.0 (R Foundation for Statistical Computing, Vienna, Austria) with RStudio (Posit Software, Boston, MA). Tables were generated using the *gtsummary* package[9]. Code is available upon request. Continuous variables are reported as median [IQR] and categorical as N (%). As appropriate, Fisher's exact test, the Kruskal-Wallis test, and the Chi-squared test were utilized for univariate statistical comparisons. P<0.05 was the prespecified threshold for significance. Code is available upon request.

### Supplementary Results

*Electrode trajectories:*

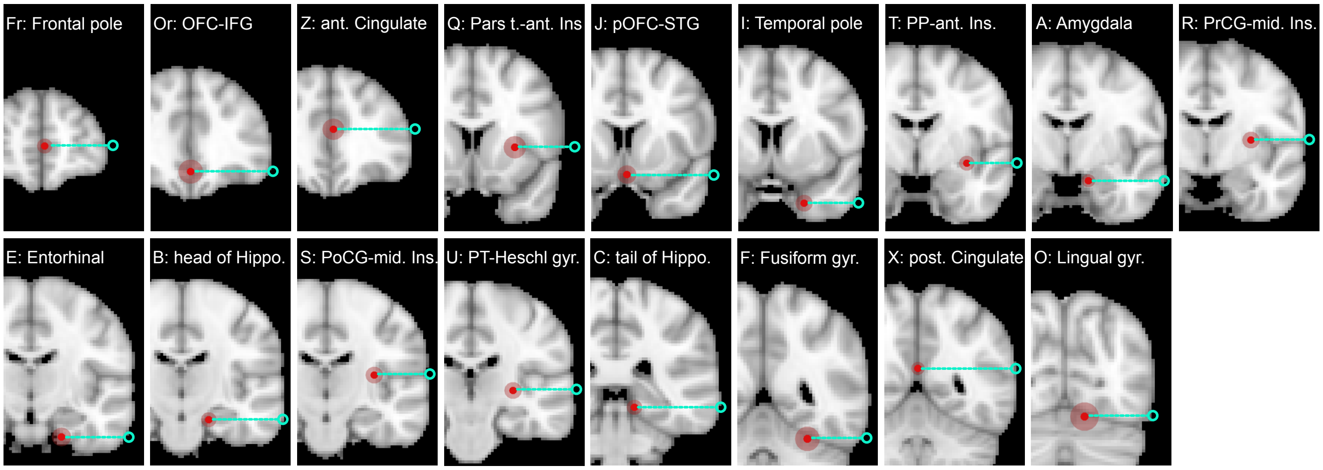


**Supplementary Figure 3: Typical electrode trajectories (coronal view)**. Bubble plots on the MNI152 T1-weighted coronal MRI demonstrating typical orthogonal implantation that permits consistent recording from both dorsolateral cortical surfaces and medial structures including the mesial temporal lobe and subcortical structures. The cyan lines and red circles represent the respective trajectories and entry points. OFC: orbitofrontal cortex, IFG: inferior frontal gyrus, ant: anterior, Pars t: Pars triangularis, Ins: insula, pOFC: posterior orbitofrontal cortex, STG: superior temporal gyrus, PP: planum polare, PrCG: precentral gyrus, Hippo: hippocampus, PoCG: postcentral gyrus, PT: planum tempolare, gyr: gyrus, post: posterior

*Case examples*:

**Case 1:** Patient is a 55-60 year-old female with epilepsy since her twenties. Semiology was reported as dizziness followed by left face twitching. In prolonged seizures, the patient was unable to talk and had focal to bilateral secondary generalization. After failing 5 anti-seizure medications, the patient continued to have focal to bilateral tonic-clonic seizures monthly. MRI showed nonspecific white matter changes over the right periventricular regions. EMU phase I data showed right frontocentral onset with rapid secondary synchronization involving bilateral frontocentral regions. SPECT injection (19 seconds) showed increased blood flow over the right insula/perisylvian regions. MEG showed increased irritability in the right posterior perisylvian regions. The proposed hypotheses were right anterior perisylvian vs. lateral temporal vs. frontal. SEEG evaluation showed interictal activity of spikes localized to the lateral STG, synchronous with the mid ventral insula, mid to posterior STG and temporal pole. Ictal EEG was notable for high amplitude spikes over the STG, temporal pole and mesial mid ventral insula followed by low voltage fast activity over the temporal pole followed by posterior STG. Direct cortical stimulation using 50Hz stimulation of anterior STG triggered the typical stereotypical seizures but not with posterior STG. Patient underwent resection of the temporal pole, anterior STG with sparing of the hippocampus, parahippocampal gyrus, and amygdala. Lateral S involving the anterior superior temporal gyrus with margin of the resection of the mid superior temporal gyrus (T electrode contacts) and resulted in Engel 1D outcome (**Supplementary Fig. 4)**.

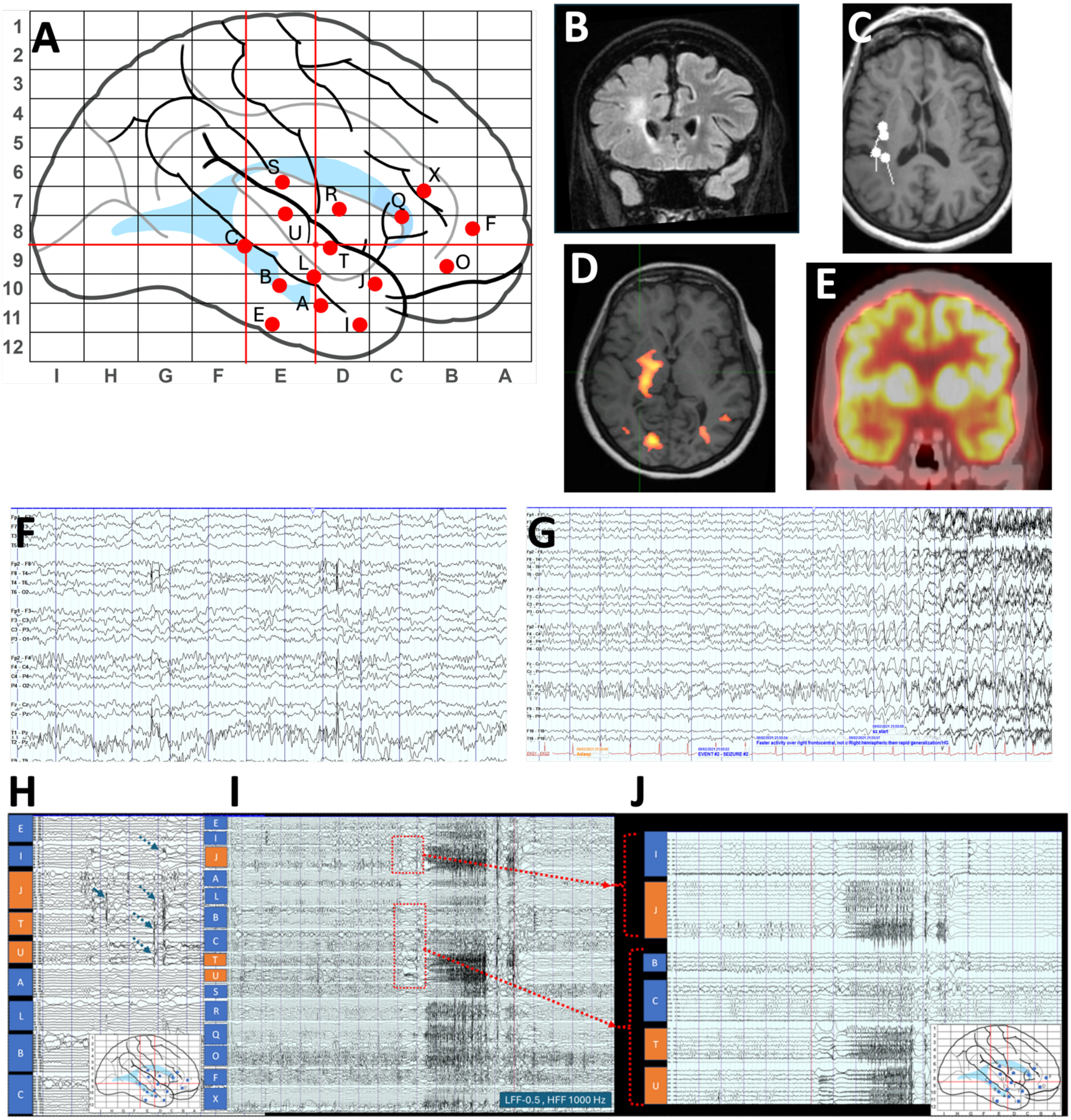


**Supplementary Figure 4: Case 1– Anterior Perisylvian Hypothesis.** A: Talairach grid demonstrating right sided implantation with standard electrode names. Note that the template anterior perisylvian implantation was expanded frontally due to the proposed hypothesis. B: Coronal T2 FLAIR image demonstrating nonspecific white matter hyperintensity in the frontal lobe. C: MEG dipoles in posterior insula and operculum overlaid over axial T1 MRI image. D: Ictal SPECT image (19 seconds after ictus) demonstrating subcortical and posterior insula uptake. E: Interictal PET with slightly decreased R temporal uptake. F: Interictal scalp video EEG with right frontotemporal sharp waves. G: Ictal scalp video EEG with typical right frontotemporal onset and bilateral generalization. H: SEEG interictal activity. SEEG evaluation showed interictal activity of spikes localized to the lateral superior temporal gyrus (J lateral electrode contacts) (Blue Arrow), at time synchronous with the mid ventral insula (T mesial contacts as well as U mid electrode contacts), mid to posterior superior temporal gyrus (T and U lateral electrode contacts) and anterior temporal regions (I mesial contacts) (blue dotted arrow). I: Ictal SEEG demonstrated huge spikes over the superior temporal gyrus, temporal pole and mesial mid ventral insula followed by low voltage fast activity over the anterior temporal pole regions (I, T and J) followed by posterior superior temporal gyrus (U lateral electrodes). J: Direct cortical stimulation using 50Hz stimulation of J lateral contacts triggered the typical stereotypical seizures but not with the U lateral contacts.

**Case 2:**
45-50 year-old female who presented with epilepsy since her twenties. She describes her seizures as an initial aura of a generalized odd feeling and panic, occasionally associated with a bad odor, progressing to a sense of altered hearing and sensation. The seizure lasts approximately 1 minute and postictally she has headache and fatigue. Her seizures occurred every 1-2 weeks despite failing adequate trials of 6 anti-seizure medications and she had a remote single generalized tonic-clonic seizure. During her phase I video EEG monitoring she was noted to initially look at her left arm and touch it with her right hand and is unable to follow commands or repeat words. She covers her eyes due to reported eye sensitivity and reports continued word finding difficulties post-ictally. Scalp EEG demonstrates left posterior temporal theta/delta slowing and frequent left posterior temporal spike/polyspikes interictally and left posterior temporal onset ictally. MRI brain demonstrated periventricular nodular heterotopia extending from the left atrium to the left temporal horn as well as atrophy of the perisylvian regions, basal temporal lobe, and cingulate gyrus on the left. Interictal MEG demonstrates left posterior temporal/parietal dipoles. Ictal SPECT (15 second injection) demonstrated increased uptake in the left posterior temporal lobe.

The proposed hypotheses were left perisylvian vs. temporal/basal/occipital vs. mesial temporal. Interictal SEEG evaluation demonstrated near continuous synchronized periodic discharges in the basal temporal and occipital aspects of the heterotopia and cortex but not in other areas, periodic discharges in the cingulate gyrus, and hippocampal head and tail spikes synchronized to discharges from the heterotopia. Ictal EEG demonstrates near synchronous onset from the PVNH and basal temporal/occipital cortices with diffuse fast activity followed by delta slowing with subsequent spread to the more anterior inferior temporal gyrus and posterior cingulate. Stimulation (1 Hz/50 Hz) in basal temporal lobe or cingulate did not elicit any seizures. After multidisciplinary discussion it was felt that both the basal temporal/occipital lobe and the cingulate gyrus could be independent EZs. She was offered resection of the basal temporal/occipital regions with potential second stage ablation of the posterior cingulate gyrus. She underwent the basal temporal/occipital resection (cortical and PVNH) without issue but never required the second stage. She achieved an Engel IA outcome and is weaning antiseizure medications (**Supplementary Figure 5**).

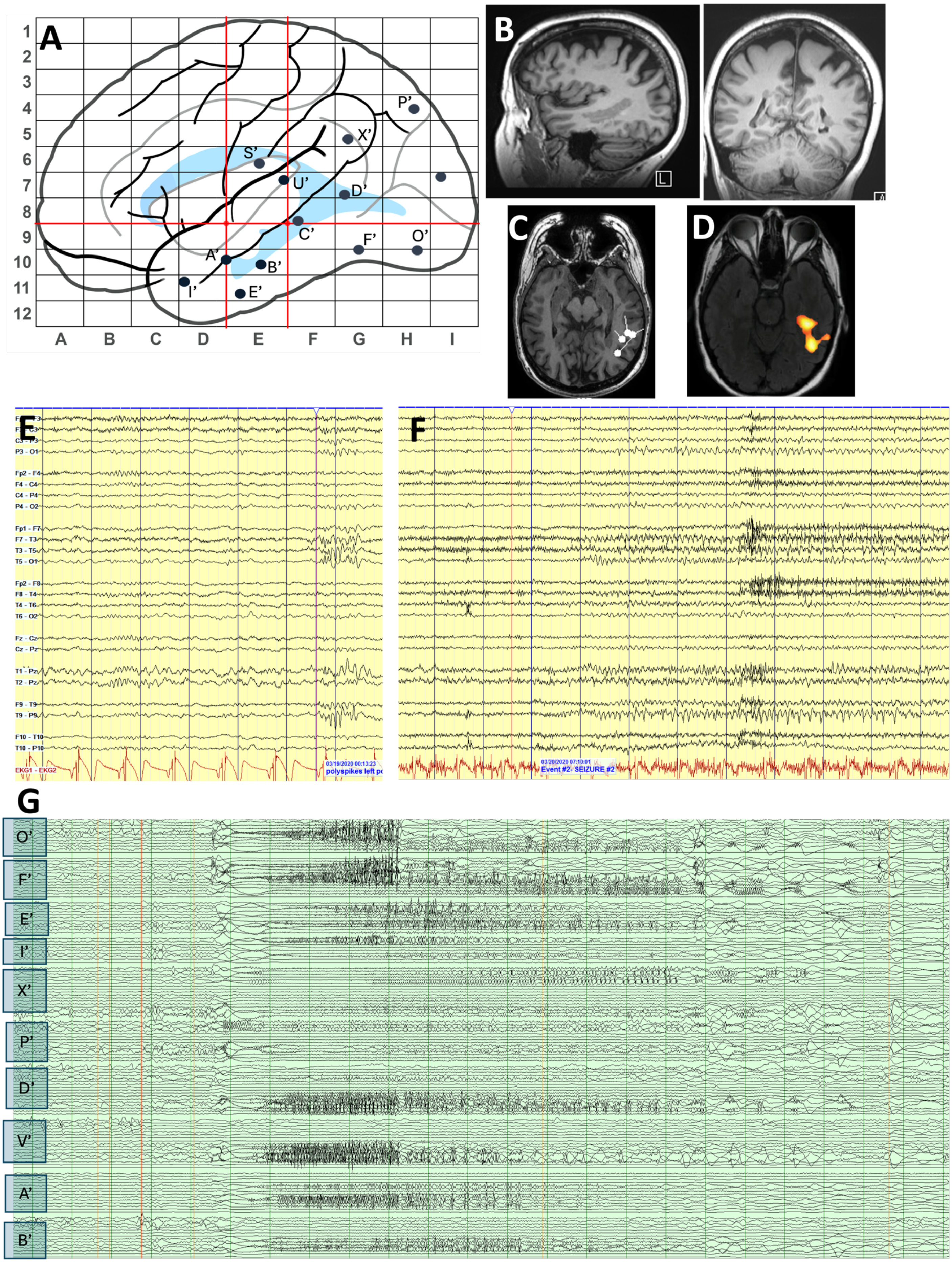
A
1
2
3
4
5
6
7
8
9
10
11
12
A B C D E F G H I
P'
X'
S'
U'
D'
C'
F'
O'
A'
B'
I'
E'
B
L
C
D
E
P3 - O1
Fp2 - F4
F4 - C4
C4 - P4
P4 - O2
Fp1 - F7
F7 - T3
T3 - T5
T5 - O1
Fp2 - F8
F8 - T4
T4 - T6
T6 - O2
Fz - Cz
Cz - Pz
T2 - Pz
F9 - T9
T9 - P9
F10 - T10
T10 - P10
EKG1 - EKG2
03/19/2020 00:13:23
polyspikes left po
F
Event #2- SEIZURE #2
G
O'
F'
E'
I'
X'
P'
D'
V'
A'
B'

**Supplementary Figure 5: Case 2 Perisylvian vs. temporal basal occipital with a periventricular nodular heterotopia**. A: Talairach grid demonstrated left-sided perisylvian vs. temporal basal occipital implantation with standard electrode names. B: Sagittal and coronal preoperative T1 MRI demonstrated extensive left periventricular nodular heterotopia extending from left atrium to temporal pole. C: Interictal MEG demonstrating left posterior temporal/basal temporal dipoles. D: Ictal SPECT (15 second injection) demonstrates left posterior temporal uptake. E: Interictal phase I scalp EEG demonstrates left posterior temporal polyspikes. F: Ictal scalp EEG demonstrates left posterior temporal onset seizures. G: Ictal SEEG demonstrates diffuse fast activity followed by delta slowing with low voltage fast activity in the posterior basal and temporal lobes (F', O', D', V' middle/lateral) spreading to the inferior temporal gyrus (E' lateral) and cingulate gyrus (X' medial) as well as STG (A', B' lateral)

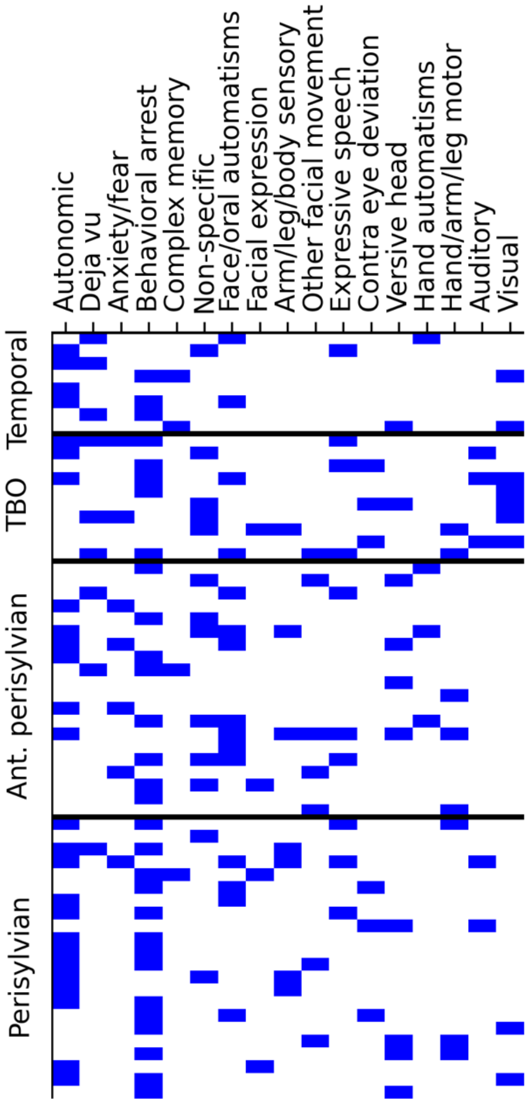


**Supplementary Figure 6: Semiological features by subject**. Each row represents an individual subject (N=60 total), organized by proposed classification. Filled cells represent observed semiology in each subject's seizures, as determined by the treating epileptologist and/or the multidisciplinary epilepsy patient management conference

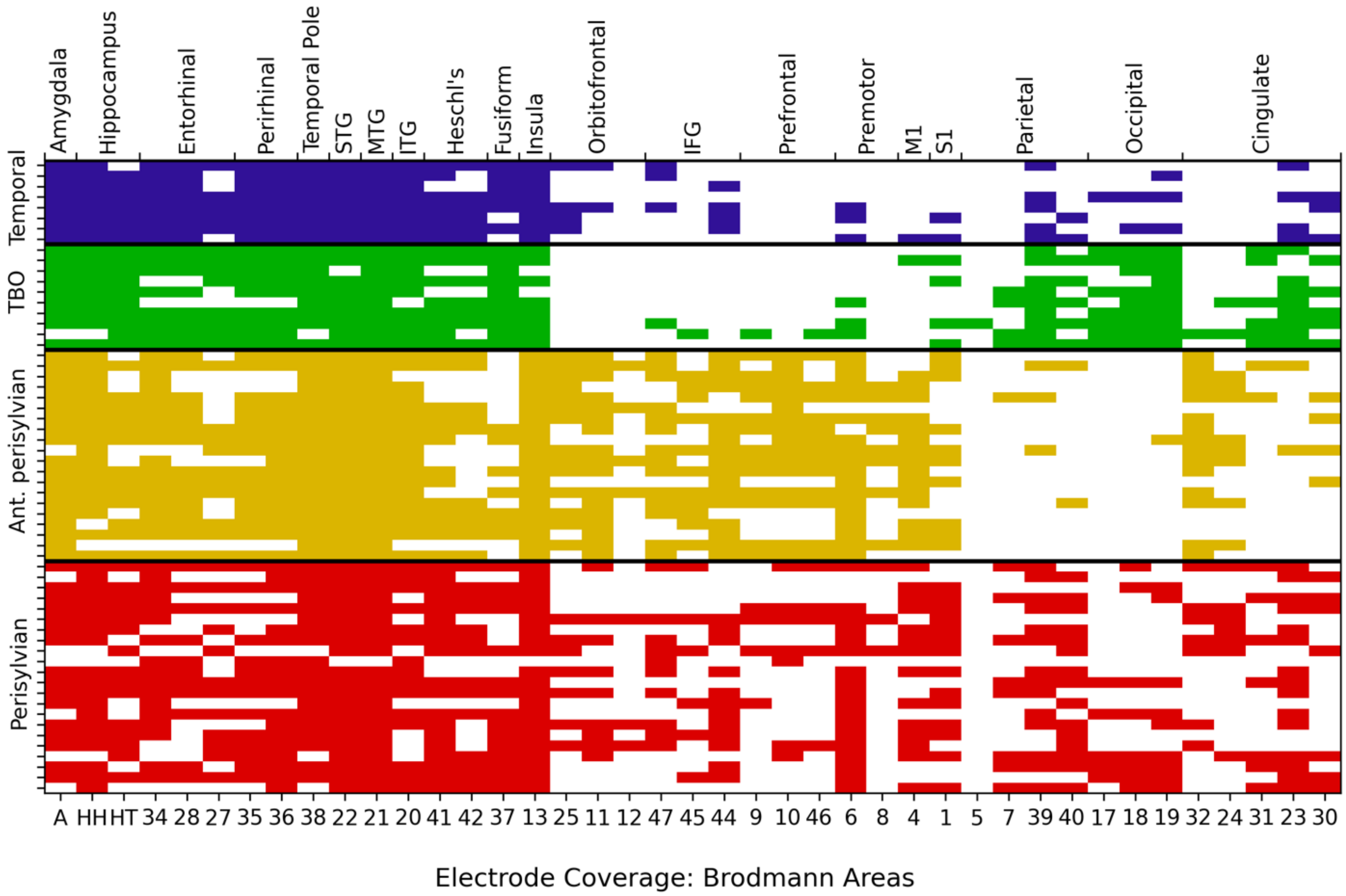


**Supplementary Figure 7: Electrode coverage by subject**. Each row represents an individual subject (N=60 total), organized by proposed classification. Filled cells represent electrode coverage in that Brodmann area (bottom x-axis) and larger region (top x-axis). Note consistent coverage of temporal regions but variable coverage of extratemporal regions by hypothesis.

**Supplementary Table 1: Electrode coordinates**

| Electrode Abbreviation | Anatomic Target | Indirect targeting coordinates [median, (IQR)] | | | | | |
|---|---|---|---|---|---|---|---|
| | | Talairach | | | MNI | | |
| | | X | Y | Z | X | Y | Z |
| A | Amygdala | ±15·7 (3·5) | -3·7 (5·4) | -12·4 (7·4) | ±17 (3) | -4 (5) | -17 (8) |
| B | Head of hippocampus | ±20·1 (4·5) | -14·3 (5·7) | -13·4 (5·6) | ±21 (4) | -15 (7) | -17 (6) |
| C | Tail of Hippocampus | ±14·5 (4·8) | -32·2 (6) | -4·6 (8·1) | ±15 (4) | -33 (7) | -7 (8) |
| E | Entorhinal cortex | ±21·9 (4·9) | -9·3 (6·5) | -24·4 (5·4) | ±22 (6) | -11 (8) | -30 (7) |
| F | Fusiform gyrus | ±30·3 (6·4) | -45·5 (7) | -17·5 (8·2) | ±31 (9) | -48 (7) | -20 (10) |
| Fr | Frontal pole | ±3·2 (3·1) | 53·5 (4·9) | 12·8 (11·2) | ±4 (4) | 59 (4) | 7 (13) |
| I | Temporal pole | ±23·5 (5·6) | 8·5 (6·9) | -26·5 (5·6) | ±25 (5) | 8 (7) | -34 (7) |
| J | Orbitofrontal cortex – anterior STG (paralimbic electrodes) | ±4·3 (4·6) | 12·6 (3·9) | -8·8 (6·6) | ±5 (6) | 13 (4) | -14 (8) |
| O | Lingual gyrus – occipital lobe | ±16·1 (14·8) | -65·6 (17·7) | -12·5 (13·7) | ±16 (13) | -69 (19) | -14 (16) |
| Or | Orbitofrontal cortex – pars orbitalis | ±3·3 (7·8) | 37·1 (8·4) | -2·85 (8·4) | ±3 (10) | 39 (10) | -9 (10) |
| Q | Pars triangularis / operculum – anterior insula | ±28·3 (6·9) | 12·3 (8·1) | 10·6 (5·1) | ±30 (8) | 15 (8) | 7 (6) |
| R | Inferior precentral gyrus – mid insula | ±32·3 (3·2) | -6·0 (8·1) | 12·2 (5·2) | ±33 (5) | -5 (8) | 10 (6) |
| S | Inferior postcentral gyrus – posterior insula | ±31·9 (5·9) | -15·6 (7·6) | 14·1 (8·2) | ±34 (5) | -15 (8) | 14 (9) |
| T | Planum polare – anterior insula | ±34·6 (4·6) | -2·7 (6·4) | -1 (6·1) | ±36 (4) | -2 (7) | -5 (7) |
| U | Planum temporale – Heschl’s gyrus – posterior insula | ±29·5 (5·2) | -21·2 (9·7) | 5·6 (5·2) | ±31 (5) | -21 (10) | 4 (6) |
| V | Cuneus – occipital lobe | ±3·4 (2·8) | -73·1 (8) | 10·6 (13·6) | ±4 (4) | -75 (8) | 13 (16) |
| Z | Anterior cingulate – frontal lobe | ±4·1 (7·8) | 35·2 (4·8) | 21·2 (8) | ±5 (8) | 39 (5) | 18 (9) |
| X | Posterior cingulate – supramarginal gyrus | ±3·0 (3·3) | -48·0 (8·7) | 18·5 (10·5) | ±3 (3) | -49 (10) | 20 (11) |

**Supplementary Table 2: Key features by hypotheses per recursive feature elimination analysis.**

| | | Temporal | Temporal Basal Occipital | Anterior perisylvian | Perisylvian | Feature Importance |
|---|---|---|---|---|---|---|
| Semiology | Behavioral arrest | 10·00% | 16·70% | 26·70% | 46·70% | 0·132 |
| | Autonomic | 15·40% | 11·50% | 23·10% | 50·00% | 0·125 |
| | Visual | 22·20% | 55·60% | 0·00% | 22·20% | 0·123 |
| | Face/oral automatisms | 13·30% | 13·30% | 46·70% | 26·70% | 0·112 |
| | Generalized body sensation | 7·70% | 30·80% | 46·20% | 15·40% | 0·101 |
| | Expressive speech | 10·00% | 30·00% | 30·00% | 30·00% | 0·092 |
| | Deja vu | 33·30% | 33·30% | 22·20% | 11·10% | 0·086 |
| | Versive head | 10·00% | 10·00% | 40·00% | 40·00% | 0·08 |
| | Anxiety/fear | 0·00% | 28·60% | 57·10% | 14·30% | 0·076 |
| | Dreamy state | 50·00% | 0·00% | 25·00% | 25·00% | 0·074 |
| Electrode Location | Prefrontal 10 | 0·00% | 0·00% | 80·00% | 31·80% | 0·128 |
| | M1 4 | 12·50% | 10·00% | 75·00% | 63·60% | 0·121 |
| | Parietal 40 | 25·00% | 50·00% | 10·00% | 68·20% | 0·116 |
| | Cingulate 23 | 50·00% | 80·00% | 10·00% | 68·20% | 0·108 |
| | IFG 44 | 50·00% | 0·00% | 85·00% | 54·50% | 0·105 |
| | Occipital 18 | 25·00% | 90·00% | 0·00% | 36·40% | 0·093 |
| | Entorhinal 28 | 100·00% | 80·00% | 75·00% | 50·00% | 0·087 |
| | Hippo· Tail | 87·50% | 100·00% | 65·00% | 68·20% | 0·084 |
| | Premotor 6 | 37·50% | 30·00% | 90·00% | 81·80% | 0·082 |
| | Orbitofrontal 11 | 25·00% | 0·00% | 85·00% | 40·90% | 0·078 |

**Supplementary Table 3: Key resection features by hypotheses per recursive feature elimination analysis.**

| | | Temporal | Temporal Basal Occipital | Anterior perisylvian | Perisylvian | Feature Importance |
|---|---|---|---|---|---|---|
| Resection | Temporal Pole 38 | 100·00% | 71·40% | 62·50% | 52·60% | 0·153 |
| | Hippocampal Tail | 14·30% | 0·00% | 37·50% | 26·30% | 0·123 |
| | Orbitofrontal 11 | 0·00% | 0·00% | 31·20% | 5·30% | 0·12 |
| | ITG 20 | 14·30% | 57·10% | 0·00% | 26·30% | 0·116 |
| | Amygdala | 100·00% | 57·10% | 56·20% | 63·20% | 0·107 |
| | Occipital 19 | 0·00% | 42·90% | 0·00% | 15·80% | 0·094 |
| | IFG 47 | 0·00% | 0·00% | 31·20% | 5·30% | 0·09 |
| | Perirhinal 35 | 71·40% | 57·10% | 50·00% | 52·60% | 0·079 |
| | Fusiform 37 | 0·00% | 42·90% | 0·00% | 21·10% | 0·063 |
| | MTG 21 | 0·00% | 14·30% | 6·20% | 21·10% | 0·055 |

**Supplementary Table 4: Factors associated with seizure freedom**

| **Characteristic** | **Not Seizure Free**, N = 20[1] | **Seizure Free**, N = 36[1] | **p-value**[2] |
|---|---|---|---|
| Sex | | | 0·5 |
| M | 10 (50%) | 21 (58%) | |
| F | 10 (50%) | 15 (42%) | |
| Age | 33 (29, 44) | 34 (26, 44) | >0·9 |
| MRI Abnormality | 12 (60%) | 25 (69%) | 0·5 |
| Number of Electrodes | 15 (13, 15) | 14 (13, 15) | 0·8 |
| Laterality | | | 0·4 |
| Right | 3 (15%) | 11 (31%) | |
| Left | 9 (45%) | 14 (39%) | |
| Bilateral | 8 (40%) | 11 (31%) | |
| Post SEEG intervention | | | <0·001 |
| Resection | 13 (65%) | 36 (100%) | |
| Neuromodulation | 7 (35%) | 0 (0%) | |
| Extent of resection | | | 0·2 |
| Temporal | 7 (54%) | 27 (75%) | |
| Extratemporal + temporal | 6 (46%) | 9 (25%) | |

[1]n (%); Median (IQR)

[2]Fisher's exact test; Pearson's Chi-squared test; Wilcoxon rank sum test

**Supplementary Table 5: Neuropsychological outcomes in resection/ablation cohort**

| Neuropsychological Test | Overall | Left | Right | p-value[1] |
|---|---|---|---|---|
| **Verbal memory (RAVLT DR)** | | | | 0·4 |
| Decline | 6 (26%) | 5 (36%) | 1 (11%) | |
| Stable | 14 (61%) | 7 (50%) | 7 (78%) | |
| Improved | 3 (13%) | 2 (14%) | 1 (11%) | |
| **Visuospatial memory (ROCF DR)** | | | | 0·4 |
| Decline | 2 (8·0%) | 2 (13%) | 0 (0%) | |
| Stable | 17 (68%) | 11 (69%) | 6 (67%) | |
| Improved | 6 (24%) | 3 (19%) | 3 (33%) | |
| **Picture Naming (BNT)** | | | | 0·05 |
| Decline | 4 (19%) | 4 (29%) | 0 (0%) | |
| Stable | 15 (71%) | 10 (71%) | 5 (71%) | |
| Improved | 2 (9·5%) | 0 (0%) | 2 (29%) | |
| **Phonemic Fluency** | | | | 0·8 |
| Decline | 0 (0%) | 0 (0%) | 0 (0%) | |
| Stable | 20 (95%) | 13 (100%) | 7 (88%) | |
| Improved | 1 (4·8%) | 0 (0%) | 1 (13%) | |
| **Semantic Fluency** | | | | 0·075 |
| Decline | 6 (29%) | 5 (38%) | 1 (13%) | |
| Stable | 10 (48%) | 7 (54%) | 3 (38%) | |
| Improved | 5 (24%) | 1 (7·7%) | 4 (50%) | |
| **Word reading (WTAR)** | | | | 0·2 |
| Decline | 2 (9·5%) | 2 (15%) | 0 (0%) | |
| Stable | 18 (86%) | 11 (85%) | 7 (88%) | |
| Improved | 1 (4·8%) | 0 (0%) | 1 (13%) | |

[1]Pearson's Chi-squared test
